# Using Relative Risk Rankings to Understand Information Differences in Multimodal Prediction Models

**DOI:** 10.1101/2025.10.30.25339162

**Authors:** Chanhwi Kim, WonJin Yoon, Hoonick Lee, Jung-Oh Lee, Majid Afshar, Jaewoo Kang, Timothy Miller

## Abstract

Recent multimodal models increasingly combine heterogeneous clinical data, yet in practice raw modalities are often replaced by expert-written summaries for convenience. However, whether such representational substitution, such as replacing medical images with their expert-written reports, preserves prognostic information has not been systematically characterized. Quantifying such gaps is essential for identifying which representational modality is most informative for predictive modeling of patient outcomes.

We investigated this question by comparing the predictive utility of chest radiographs (CXRs) and their paired radiology report text for 30-day post-discharge mortality prediction using vision-language models (VLMs). Using the discharge note summary as global clinical context at discharge, we augmented it with either the latest pre-discharge CXR or the corresponding radiology report. On a linked MIMIC-IV/MIMIC-CXR subset with paired inputs (n = 1,360), the discharge-note-plus-CXR model achieved the best performance (AUROC = 0.864), compared with the discharge-note-only model (AUROC = 0.831) and the discharge-note-plus-report model (AUROC = 0.813).

To quantify the effect of modality substitution on predictive behavior, we measured Kendall’s *τ*-based distances between predicted risk rankings. Inter-modality distances exceeded intra-modality distances, indicating that replacing CXRs with reports changes risk prioritization rather than merely reducing overall discrimination. Post hoc review by a radiologist further suggested that reports, as clinically oriented summaries, may not exhaustively document visually available prognostic cues that could contribute to model risk stratification. Overall, our findings suggest that expert-written reports may be imperfect prognostic proxies for raw images and that representational substitution should be evaluated using both discrimination and ranking agreement.

## 1. Introduction

Predictive models in healthcare increasingly integrate heterogeneous data modalities, yet clinical information differs not only in content but also in representation (Li, Wang, Law, Murali and Pandey, 2022). The same underlying clinical state may be expressed as a raw medical image or as an expert-written report, where the latter is a selective and clinically motivated summary rather than a lossless transcription of all image-visible findings (Waite, Scott, Gale, Fuchs, Kolla and Reede, 2017; Jain, Smit, Truong, Nguyen, Huynh, Jain, Young, Ng, Lungren and Rajpurkar, 2021; Nobel, van Geel and Robben, 2022). In practice, such expert-written summaries are often used as convenient substitutes for raw data (Jain et al., 2021; Lin, Yang, Yin, Tang, Chen, Xu, Zhu, Gao, Liu, Liu et al., 2024). However, whether this kind of representational substitution preserves prognostic information remains insufficiently characterized.

This question is particularly relevant for post-discharge risk assessment after intensive care. Patients discharged from the ICU remain at elevated risk of death and readmission, making early identification of high-risk patients clinically important (Afessa and Keegan, 2007; Ghassemi, Naumann, Doshi-Velez, Brimmer, Joshi, Rumshisky and Szolovits, 2014; Rajkomar, Oren, Chen, Dai, Hajaj, Hardt, Liu, Liu, Marcus, Sun et al., 2018). In particular, mortality prediction has received major attention from researchers because patients discharged from the ICU remain at elevated risk of death and readmission (Grnarova, Schmidt, Hyland and Eickhoff, 2016; Sushil, Šuster, Luyckx and Daelemans, 2018; Purushotham, Meng, Che and Liu, 2018; Lin et al., 2024). Electronic health records (EHRs), which contain both structured and unstructured data, have been widely utilized to address these risks. In particular, structured EHR data, such as demographics, vital signs, and laboratory results, has been successfully employed for mortality prediction (Naved, Siddiqui and Khan, 2011; McGilvray, Heaton, Guo, Masood, Cupps, Damiano, Pasque and Foraker, 2022; Choi, Kim, Choi, Jung, Choi, Cho and Jeong, 2022). Recent advances in natural language processing (NLP) and large language models (LLMs) have facilitated the extraction of prognostic signals from unstructured clinical narratives (Tran and Lee, 2018; Hashir and Sawhney, 2020; Mahbub, Srinivasan, Danciu, Peluso, Begoli, Tamang and Peterson, 2022). Among these texts, discharge notes are especially valuable for mortality prediction, as they provide comprehensive clinical information essential for post-discharge risk assessment (Luo and Rumshisky, 2017). Advanced clinical NLP models can effectively process long textual contexts and capture clinically meaningful information that is often absent from structured data (Huang, Altosaar and Ranganath, 2019; Yang, Chen, PourNejatian, Shin, Smith, Parisien, Compas, Martin, Costa, Flores et al., 2022; Luo, Sun, Xia, Qin, Zhang, Poon and Liu, 2022; Gana Castillo, Figueroa Mandujano, Allende-Cid, Nand, Zamora and Ramos, 2024). Building on these advances, models that jointly leverage structured and unstructured data have achieved strong performance in predicting patient outcomes (Garriga, Buda, Guerreiro, Iglesias, Aguerri and Matić, 2023; Ye, Hai, Song and Wang, 2024).

Beyond structured and unstructured EHR data, chest radiographs (CXRs) are also widely used to assess cardiopulmonary patient status (Lu, Ivanov, Mayrhofer, Hosny, Aerts and Hoffmann, 2019; Sanida, Sanida, Sideris and Dasygenis, 2024; Weiss, Raghu, Paruchuri, Zinzuwadia, Natarajan, Aerts and Lu, 2024). Radiology reports, in turn, provide expert-written summaries of major findings or changes in a patient’s thoracic status and are therefore often more accessible for downstream modeling. Consequently, CXRs and radiology reports have been integrated with structured EHR data to predict patient outcomes using machine learning and deep learning approaches (Mohsen, Ali, El Hajj and Shah, 2022; Soenksen, Ma, Zeng, Boussioux, Villalobos Carballo, Na, Wiberg, Li, Fuentes and Bertsimas, 2022; Lin, Wang, Ding, Zhao, Wang and Peng, 2025). Some studies also incorporated unstructured clinical text, although most approaches have continued to rely on conventional deep learning pipelines (Lin, Wang, Ding, Zhao, Wang and Peng, 2021; Wang, Yin and Zhang, 2024). These efforts demonstrated that CXRs contain prognostic information beyond that available in both structured and unstructured EHR data. However, these studies have largely focused on whether adding each modality improves aggregate predictive performance, rather than on whether replacing one representation with another preserves admission-level predictive behavior. Accordingly, it remains unclear whether radiology reports can serve as prognostic substitutes for raw CXR images (JPEG format) when both are evaluated under the same clinical context. To the best of our knowledge, no prior research has applied vision–language models (VLMs) to CXRs alongside discharge notes for mortality prediction while systematically comparing the ranking behavior when substituting CXR representations.

In this study, we examine whether representational modality affects 30-day post-discharge mortality prediction. Using the discharge note as shared clinical context at discharge, we compare models augmented with either the latest predischarge CXR or its paired radiology report. Beyond aggregate discrimination, we also assess whether these alternative representations lead to different patient risk rankings, since two models with similar overall performance may still prioritize different patients as high risk.

Accordingly, we address three research questions:

1. Do CXR-augmented and report-augmented models, conditioned on the same discharge note context, produce systematically different discriminative performance and mortality risk rankings?
2. How large are these modality-associated ranking differences relative to within-modality variation, as quantified by rank-based agreement measures?
3. What qualitative differences between paired CXRs and radiology reports may help contextualize these ranking disagreements?

We define 30-day post-discharge mortality as the primary endpoint and evaluate discriminative performance using the area under the receiver operating characteristic curve (AUROC) and the area under the precision–recall curve (AUPRC). Because discrimination alone does not capture whether models prioritize the same patients as high risk, our primary analysis focuses on modality-associated differences in predicted risk ordering using Kendall’s *τ*-based distance. It enables a rank-aware comparison between CXR-augmented and report-augmented predictors under the same discharge-note context. Finally, we contextualize observed disagreements through post hoc review of paired CXRs and reports, examining cases in which image-visible details relevant to risk stratification are not explicitly documented in the corresponding report. Together, these analyses characterize how representational choices affect both predictive performance and admission-level prioritization in multimodal clinical prediction.

## 2. Related Work

### 2.1. Mortality Prediction from EHR and Clinical Text

Early work on ICU mortality prediction primarily relied on structured EHR variables such as vital signs, laboratory measurements, and demographics (Johnson, Pollard, Shen, Lehman, Feng, Ghassemi, Moody, Szolovits, Anthony Celi and Mark, 2016; Harutyunyan, Khachatrian, Kale, Ver Steeg and Galstyan, 2019). Although these data provide objective summaries of physiological status, they are often sparse, irregularly sampled and may not fully capture contextual information documented in clinical narratives. To address this limitation, subsequent studies increasingly incorporated unstructured clinical text, including progress notes or discharge summaries, which contain dense descriptions of patient trajectory, treatments, and clinician assessment. Initial approaches applied recurrent and convolutional architectures to model clinical notes (Shickel, Tighe, Bihorac and Rashidi, 2017). More recently, Transformer-based language models such as ClinicalBERT and BioBERT have improved extraction of prognostic information from long-form notes by modeling contextual semantics and long-range dependencies (Alsentzer, Murphy, Boag, Weng, Jindi, Naumann and McDermott, 2019; Lee, Yoon, Kim, Kim, Kim, So and Kang, 2020). Large language models have further expanded text-based modeling, including applications to both outcome prediction and clinical summarization (Yang et al., 2022; Nazi and Peng, 2024). However, text-only predictors remain constrained by what is explicitly documented and therefore may miss complementary information observable in other modalities such as medical imaging.

### 2.2. Prognostic Value of Chest Radiographs and Radiology Reports

Chest radiographs (CXRs) are routinely used in critical care and provide visual evidence of thoracic pathology, device placement, and overall cardiopulmonary status (Ganapathy, Adhikari, Spiegelman and Scales, 2012). Deep learning models, particularly CNN-based approaches, have achieved strong performance on CXR interpretation tasks such as detecting pneumonia and pneumothorax (Rajpurkar, Irvin, Zhu, Yang, Mehta, Duan, Ding, Bagul, Langlotz, Shpanskaya et al., 2017; Irvin, Rajpurkar, Ko, Yu, Ciurea-Ilcus, Chute, Marklund, Haghgoo, Ball, Shpanskaya et al., 2019). Prior work has suggested that chest radiographs may provide prognostic information relevant to mortality prediction (Lu et al., 2019). Radiology reports translate visual findings into clinically actionable expert-written summaries of these examinations and are widely used in clinical workflows. In vision language modeling, radiology reports also serve as supervision signals for learning joint image-text representations and for automated report generation (Zhang, Jiang, Miura, Manning and Langlotz, 2022; Wang, Wu, Agarwal and Sun, 2022). In downstream clinical modeling, these reports are sometimes used as practical textual proxies for images, but the extent to which reports preserve information relevant to prognostic tasks can vary (Smit, Jain, Rajpurkar, Pareek, Ng and Lungren, 2020; Tu, Azizi, Driess, Schaekermann, Amin, Chang, Carroll, Lau, Tanno, Ktena et al., 2024). Thus, radiology reports are selective clinical summaries, motivating the question of whether they fully preserve prognostic information present in raw images.

Existing multimodal studies have shown that CXRs and radiology reports can each improve clinical outcome prediction when combined with other data sources, but they have largely focused on aggregate performance gains (Lin et al., 2025).We argue that, while performance improvements are valuable, it is important to start to gain a deeper understanding of where these improvements come from by learning what kind of information each representation type effectively models. For a given prediction task, one would like to know to what extent different modalities may have complementary information, and this work describes methods that produce that information. We demonstrate the use of those tools to study a specific instance of that question: do radiology reports preserve the prognostic information of raw CXRs for predicting post-discharge mortality? This gap motivates our study, which directly compares CXR- and report-based augmentation for post-discharge mortality prediction and evaluates not only discrimination but also rank-level agreement in patient risk prioritization.

## 3. Methods

We compare prediction behavior between CXR-augmented and report-augmented models under the same discharge-note context. To quantify modality-associated differences, we measure disagreement in predicted patient risk rankings using Kendall’s *τ*-based distance. This rank-based analysis complements standard discrimination metrics by assessing whether different input modalities lead to different admission-level risk prioritization.

### 3.1. Study Objective and Task Definition

The primary task in this study is 30-day post-discharge mortality prediction for ICU patients, following the definition of Yoon, Chen, Gao, Zhao, Dligach, Bitterman, Afshar and Miller (2025a). All admissions in the cohort included an ICU stay during the indexed hospitalization. It is formulated as a binary classification problem where the objective is to predict whether a patient will die within 30 days after discharge (*y* ∈ {0,1}).

### 3.2. Rank-based Behavioral Disagreement

We evaluated predictive performance using the area under the receiver operating characteristic curve (AUROC) and the area under the precision–recall curve (AUPRC). AUROC is a threshold-independent metric that can be interpreted as the probability that a randomly selected positive example receives a higher score than a randomly selected negative example. AUPRC summarizes the precision–recall trade-off across decision thresholds and is often more informative than AUROC in the presence of class imbalance. Because the positive class prevalence in our cohort is below 10%, we also report AUPRC as an important complementary metric to AUROC.

To quantify agreement in admission-level risk prioritization across models, we additionally measured rank correlation using Kendall’s *τ* with tie correction (*τ*_*b*_) (Kendall, 1945). Let 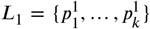 and 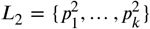 denote the predicted probabilities from two models *M*_*1*_ and *M*_*2*_ on the same set of *k* examples, and let *R*_*1*_ and *R*_*2*_ be the corresponding rank orderings. Define

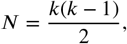

*P =* #{concordant pairs},

*Q =* #{discordant pairs},

*T =* #{tied pairs in *R*_1_},

*U =* #{tied pairs in *R*_*2*_}.

The tie-corrected Kendall’s tau is

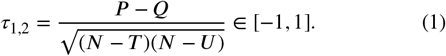

We then define the *τ*-based distance as

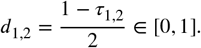

where *d*_1,2_ - 0 indicates identical rankings, *d*_1,2_ - 0.5 represents random correlation, and *d*_1,2_ = 1 signifies maximal disagreement, achieved when one ranking is the complete reverse of the other (Yoon, Ren, Thomas, Kim, Savova, Hall and Miller, 2025b).

We computed two types of distances:

- *Intra-modality distances:* distances between models trained under the same input representation (e.g., discharge note + CXR).
- *Inter-modality distances:* distances between models trained under different representations (e.g., discharge note + CXR vs. discharge note + report).

To assess the effect of random initialization during finetuning, we trained *n* models *M*_*1*_, …, *M*_*n*_ with different seeds (*n* = 5). For each pair of models *(M*_*i*_, *M*_*j*_), we computed the *τ-based distance d*_*i,j*_ between their predicted rankings on the same evaluation set.

### 3.3. Post hoc analysis

We conducted two post hoc qualitative analyses involving chest radiographs. First, a radiology expert was provided with randomly selected paired CXRs and radiology reports and asked to identify image-visible findings that could be relevant to mortality but were not explicitly described in the corresponding reports. We then grouped these observations into categories based on their clinical relevance.

Second, to qualitatively assess model attention patterns, a radiologist reviewed two image-only visualizations derived from the fine-tuned VLM for randomly selected CXRs. One was an activation-based map constructed from the patchwise vector norm of the Layer 0 LayerNorm output of the vision encoder, projected onto the image as a smoothed heatmap. The other was an occlusion-based attribution map computed over the model’s inferred patch grid. Local regions were sequentially masked using a sliding occlusion procedure, and the change in the predicted mortality logit at each location was visualized as a color-coded map. The red regions indicate areas whose occlusion increases predicted survival probability (i.e., regions associated with higher mortality risk when visible), and blue regions indicate areas whose occlusion decreases predicted survival probability (i.e., regions associated with lower mortality risk when visible). The radiologist was asked to assess whether the highlighted regions appeared clinically meaningful and whether they emphasized findings visible on the image but not explicitly described in the corresponding radiology report. The vision encoder processes 560 x 560 image tiles with a patch size of 14, corresponding to a 40 x 40 patch grid per tile.

## 4. Experiments

### 4.1. Datasets and Cohort Construction

In this study, we constructed a cohort by integrating textual and imaging data from two publicly available sources. The discharge notes were obtained from the LCD benchmark dataset (Yoon et al., 2025a), which is derived from MIMIC-IV (Johnson, Bulgarelli, Shen, Gayles, Shammout, Horng, Pollard, Hao, Moody, Gow et al., 2023), while the corresponding radiology reports and CXRs were retrieved from MIMIC-IV-CXR (Johnson, Lungren, Peng, Lu, Mark, Berkowitz and Horng, 2019; Johnson, Pollard, Mark, Berkowitz and Horng, 2024).

The textual inputs consist of de-identified discharge notes and radiology reports associated with hospital admissions at the Beth Israel Deaconess Medical Center (2008–2019), linked to out-of-hospital mortality outcomes from the Massachusetts State Registry of Vital Records and Statistics. LCD Benchmark (Yoon et al., 2025a) constructs its cohort by selecting only admissions that included an ICU stay and linking each discharge to out-of-hospital mortality after discharge. The unit of prediction is the admission, but the splits are partitioned at the patient level so that no patient appears in more than one split. Then, we restricted the cohort to patients who had at least one CXR examination during their stay. The final dataset contained 6,345 admissions in the training set, 1,367 admissions in the development set, and 1,360 admissions in the test set. The positive sample ratio in each split is 4.6%,6.1%, and 4.4% respectively. Since the original discharge notes are too long to finetune LLMs with our hardware, we first summarize the discharge notes. We summarized the original discharge note using the same model (Llama-3.2-11B-Vision-Instruct). Appendix D summarizes the distribution of studies and CXRs per admission, and Appendix E provides statistics on discharge note token lengths.

When multiple CXR studies were available for each admission, we selected the most recent and regularized image that best represented the patient’s clinical status. Ideally, leveraging all available CXRs would better capture longitudinal changes in patient status. However, this approach was limited by (i) the large variability in the number of chest radiographs (ranging from 1 to more than 30) and (ii) the lack of validated models capable of handling long, interleaved multi-image sequences. To address this, a heuristic decision tree developed with physician input was adopted. Each image was assigned a priority score based on procedure type, view, and orientation. This prioritization scheme ensured that the model received images captured under comparable conditions, thereby facilitating consistent feature extraction and reflected typical clinical practice. Finally, we chronologically sorted images in the highest-priority group and selected the most recent image prior to discharge to provide a consistent, discharge-proximal representation of the patient’s condition. The pseudocode for the selection procedure is provided in Appendix G.

### 4.2. Model training

We adopted a vision–language model to incorporate multimodal inputs, including chest radiographs and radiology reports. We evaluated multiple candidate vision–language models and selected Llama-3.2-11B-Vision-Instruct as the base architecture for all subsequent experiments (Appendix A). We employed Low-Rank Adaptation (LoRA) for parameter-efficient fine-tuning (Shen, Wallis, Allen-Zhu, Li, Wang et al.). LoRA introduces trainable low-rank update matrices into selected layers while keeping the pretrained backbone weights frozen, thereby reducing the number of trainable parameters and the overall memory footprint. We optimized the LoRA parameters jointly with the multimodal projector and a task-specific classification head attached to the final hidden representation. Training was performed on the training split, hyperparameters were selected on the development split, and final results were reported on the held-out test split. We applied identical preprocessing across all splits to ensure consistency. During evaluation, we set the temperature to 0 for deterministic prediction. Detailed hyperparameter settings are provided in Appendix F.

## 5. Results

### 5.1. Main Discrimination Results

Table 1 summarizes AUROC and AUPRC for 30-day post-discharge mortality prediction across model families and input configurations. We report results for two text-only reference baselines–an SVM (Cortes and Vapnik, 1995) with TF–IDF bag-of-words features and ModernBERT (Warner, Chaffin, Clavié, Weller, Hallström, Taghadouini, Gallagher, Biswas, Ladhak, Aarsen et al., 2025)–and for the Llama-3.2-11B-Vision-Instruct vision–language model (VLM) (Meta, 2024). Because the SVM and ModernBERT cannot directly process images, they are evaluated under DN (discharge note) and DN+RR (discharge note + radiology report), whereas the VLM is evaluated under DN, DN+RR, and DN+CXR (discharge note + chest radiograph).

**Table 1.** Discrimination performance (AUROC, AUPRC) for 30-day post-discharge mortality prediction under different input configurations and model families. DN: discharge note; RR: radiology report; CXR: chest radiograph; SVM: support vector machine. Rows correspond to progressively richer clinical context (– *Radiology Info, Discharge Summary, + Demographics)*. For LLaMA 3.2-11B, * and ** denote statistically significant differences in AUROC relative to **DN+CXR** within the same row (DeLong’s test; *p <* 0.05 and *p <* 0.01, respectively). Best results within each row are shown in bold.

| Input Configuration | SVM |  |  |  | ModernBERT |  |  |  | LLaMA 3.2-11B |  |  |  |  |  |
| --- | --- | --- | --- | --- | --- | --- | --- | --- | --- | --- | --- | --- | --- | --- |
|  | DN |  | DN+RR |  | DN |  | DN+RR |  | DN |  | DN+RR |  | DN+CXR |  |
|  | AUROC | AUPRC | AUROC | AUPRC | AUROC | AUPRC | AUROC | AUPRC | AUROC | AUPRC | AUROC | AUPRC | AUROC | AUPRC |
| – Radiology Info | 0.753 | 0.134 | 0.767 | 0.125 | 0.761 | 0.175 | 0.740 | 0.165 | 0.825* | 0.261 | 0.803** | 0.231 | **0.850** | **0.279** |
| Discharge summary | 0.784 | 0.206 | 0.797 | 0.206 | 0.792 | 0.237 | 0.805 | 0.211 | 0.831* | 0.307 | 0.813** | 0.278 | **0.864** | **0.320** |
| + Demographics | 0.802 | 0.194 | 0.813 | 0.217 | 0.808 | 0.231 | 0.817 | 0.238 | 0.841* | 0.279 | 0.823** | 0.284 | **0.875** | **0.327** |

Across the text-only baselines, ModernBERT consistently outperformed the SVM, while the effect of adding radiology reports was mixed across summary variants. These reference results indicate that additional report text does not uniformly improve mortality prediction even within text-only modeling. However, our primary comparison concerns the VLM setting, where the discharge note provides shared clinical context and the added modality differs between radiology reports and raw CXRs. Within the VLM, DN+CXR achieved the strongest performance across all summary configurations, outperforming both DN and DN+RR in AU-ROC and AUPRC. By contrast, adding radiology reports to the discharge note produced limited or inconsistent gains relative to DN alone and often reduced performance. These results indicate that raw CXRs provide prognostic information that is not fully preserved when the same examination is represented only through its paired radiology report.

This pattern was consistent across input variants and was supported by DeLong’s tests for correlated ROC curves, which showed statistically significant AUROC improvements of DN+CXR over the corresponding DN and DN+RR configurations. These results suggest that, under shared discharge-note context, radiology reports do not serve as flawless prognostic substitutes for raw CXRs in this task.

Figure 1 further illustrates this pattern through the ROC (top) and precision–recall (bottom) curves for the VLM. Across all three discharge-note summary configurations, DN+CXR shows the most favorable discrimination and precision-recall trade-off, whereas DN+RR provides little benefit over DN and is often slightly worse. Under the demographics-augmented summary setting, this advantage is especially consistent across the full recall range. Appendix B further supports this tendency at the admission level, showing that incorporating CXRs shifts predicted logodds in a more outcome-consistent direction compared to adding radiology reports. Overall, these results suggest that raw imaging contributes complementary predictive signal beyond text-only inputs, whereas radiology reports do not fully substitute for the prognostic information available in CXRs.

**Figure 1.**
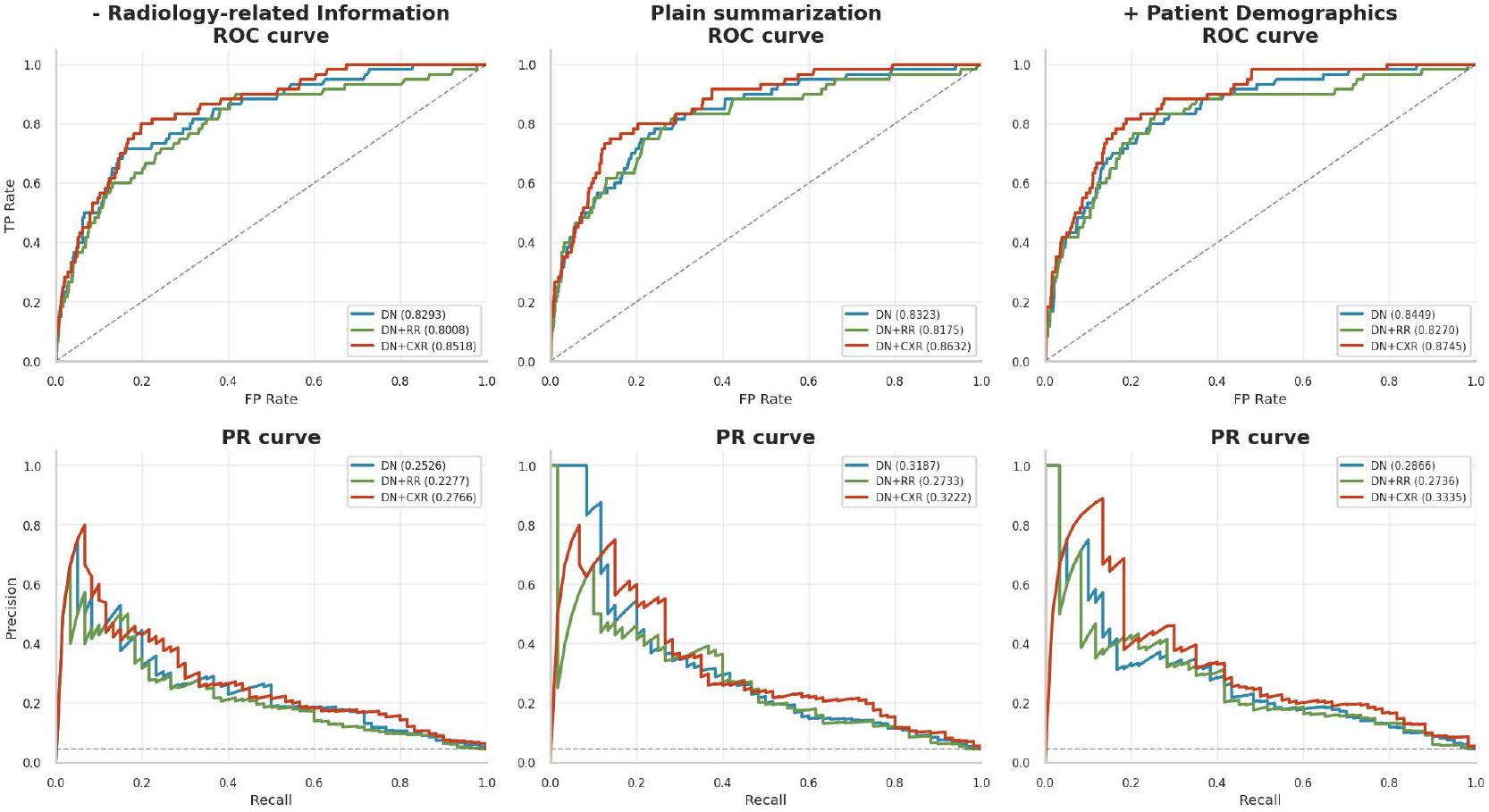
Receiver operating characteristic (ROC; top) and precision–recall (PR; bottom) curves for 30-day post-discharge mortality prediction: summaries with radiology-related content removed (left), discharge summaries (middle), and summaries augmented with patient demographics (age, race; right).

### 5.2. Disagreement in patient risk ranking

Figure 2 quantifies modality-driven differences in admission level risk ordering using Kendall’s *τ*-based distance. As a reference, grey violins show distances comparing with random ranking baseline, clustering near 0.5 as expected. Across both panels, *Intra* comparisons are consistently lower than *Inter* comparisons, indicating that models trained with the same input configuration (but different random seeds) learn broadly similar ranking behavior, whereas changing the modality leads to larger shifts in risk stratification.

**Figure 2.**
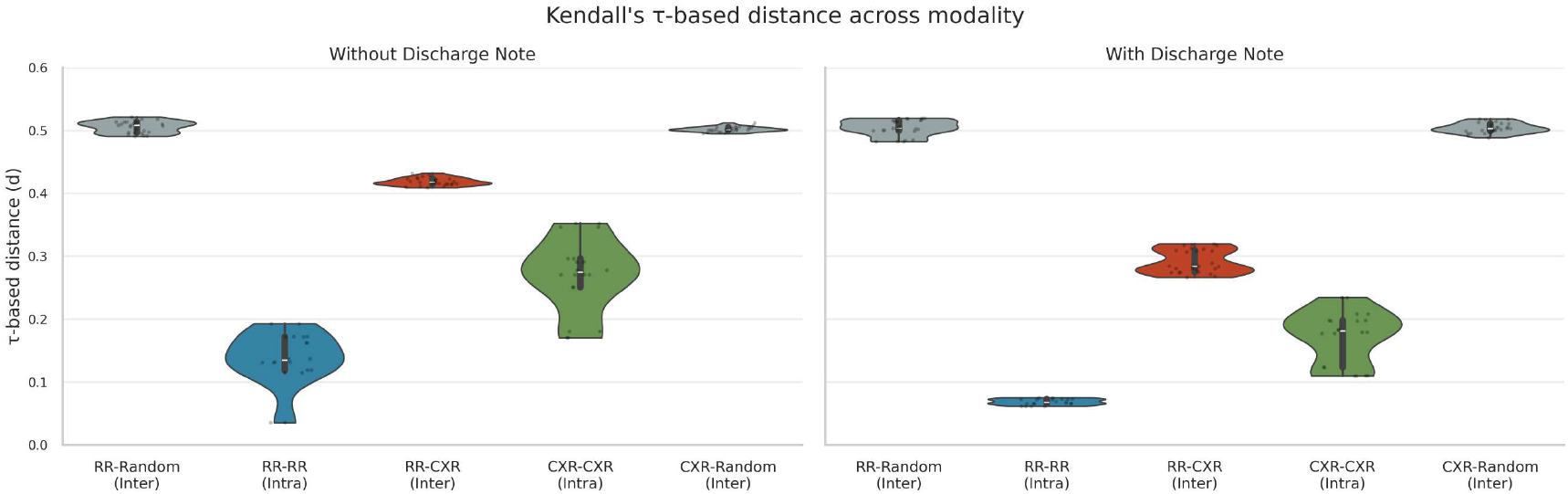
Distribution of Kendall’s *τ*-based distances between admission-level risk rankings across modalities. The left part reports comparisons between models trained with radiology-only inputs, while the right panel reports comparisons between models that share the same discharge-note context. “Intra” denotes comparisons between models trained under the same input configuration, and “Inter” denotes comparisons between models trained under different configurations. Grey violins show the *τ*-based distance against a random ranking baseline. Lower *τ*-based distance indicates more similar admission-level ranking behavior.

In the radiology-only input setting (left), CXR-based models exhibit higher *Intra* distances (0.18–0.35) than RR-based models (0.05–0.20). By contrast, *Inter* comparisons between RR and CXR models yield substantially larger distances (above 0.4), highlighting pronounced disagreement in admission-level ordering across modalities. When discharge notes are shared (right), overall distances decrease, consistent with the shared DN context constraining model behavior; nevertheless, the *Inter* distances between DN+RR and DN+CXR (0.28–0.32) remain clearly above both *Intra* baselines, indicating that the choice of augmentation (report vs. image) still induces meaningful ranking differences.

### 5.3. Radiologist Post hoc Review Findings

Table 2 presents the results of a radiologist’s post hoc review of 40 randomly selected pairs of CXRs and their corresponding radiology reports. The radiologist identified findings that were visible on the CXRs but not explicitly documented in the reports. These findings were categorized into anatomical and clinical groups for clarity. In the lung/diaphragm category, various unreported findings were observed, including pleural effusion, atelectasis, and emphysema. Many of these findings were documented only as “stable findings” without specific description. Additional omissions were noted across several other categories: skeletal degeneration (e.g., diffuse bony degeneration, osteopenia), mediastinal findings (e.g., aortic calcification, diverticulum with calcification), cardiomegaly, post-surgical findings (e.g., sternotomy wires, surgical clips), medical devices (e.g., NG tube, ET tube, EKG leads), and body habitus indicators (e.g., obesity, senile changes). While most unreported findings represent common age-related changes or routine medical devices that are frequently omitted from reports, some of these omissions were clinically relevant.

**Table 2.** Categorized image-visible findings identified in a random subset of 40 paired CXR–report cases. These categories summarize abnormalities that were visible on the CXRs and judged by an expert radiologist to be potentially relevant, but were not explicitly documented in the corresponding official radiology reports. The omitted findings ranged from potentially clinically informative abnormalities to more routine or commonly underreported observations, such as age-related or degenerative changes, support devices, and findings described only implicitly or as stable without further specification.

| Category | Findings not documented in the radiology report |
| --- | --- |
| Lung / Diaphragm | Coarse lung parenchyma, diaphragmatic flattening, retrocardiac opacity, mild atelectasis, emphysema, pleural effusion |
| Mediastinum | Aortic calcification/atherosclerosis, unfolded aorta, esophageal diverticulum with calcification, pneumo-mediastinum |
| Skeletal degeneration | Diffuse bony degeneration, shoulder arthrosis, costochondral junction calcification, osteopenia |
| Heart | Cardiomegaly |
| Post-surgical findings | Sternotomy wire, surgical clips (CABG) |
| Devices | NG tube, ET tube, chest tube, CVC, EKG leads/electrodes, facial/air mask |
| Body habitus | Senile changes (skin wrinkling), young skeletal appearance, obesity, cachexia |
| Other | Poor positioning |

In the visualization review, both map types highlight peripheral soft tissues, such as the shoulders, neck, and abdomen, rather than the lung fields, which are typically considered the primary region of clinical interest on chest radiographs. Activation-based maps produced relatively sharper highlights, concentrating on the shoulders, neck, and mediastinal area. In contrast, occlusion-based maps showed broader, more diffuse highlighting across soft tissues, without a clearly discernible anatomical pattern. Figure 3 provides a representative example of both map types. The activation map highlights regions that potentially correspond to pathologic findings, such as pleural effusion and cardiomegaly. It also extends to normal abdominal soft tissues, as well as the right shoulder and neck. This pattern was commonly observed and does not correspond to structures routinely described in radiology reports. The occlusion map for this sample contains only blue regions (areas associated with lower mortality risk when obscured). These regions encompass both pathologic findings, such as pleural effusion and cardiomegaly, and non-pathologic diffuse soft tissue. Overall, these reviews did not reveal a consistent, clinically interpretable pattern linking the highlighted regions to predicted mortality risk.

**Figure 3.**
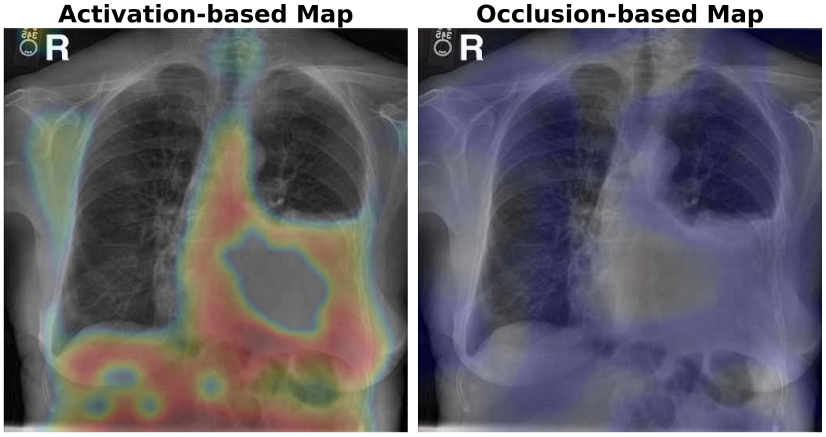
Illustrative examples of post hoc visual analysis. The left image (Activation-based Map) shows the patch-wise activation norm from the Layer 0 LayerNorm output, where warmer colors indicate higher feature activation. The right image (Occlusion-based Map) shows the occlusion sensitivity map, where blue regions indicate areas whose occlusion increases predicted mortality probability, and red regions indicate areas whose occlusion decreases predicted mortality probability.

## 6. Discussion

This study examined whether replacing raw chest radiographs (CXRs) with their paired radiology reports preserves prognostic information for 30-day post-discharge mortality prediction under a shared discharge note context. Across discharge note variants, augmenting discharge notes with CXRs consistently achieved stronger discrimination than either discharge notes alone or discharge notes augmented with radiology reports. These findings suggest that, in this cohort and modeling setup, raw imaging contains prognostic information that is not fully preserved when the same examination is represented only through its expert-written report.

This difference was reflected not only in aggregate performance but also in risk stratification at the admission level. The rank-based analysis (Fig. 2) showed that CXR-augmented and report-augmented models produced different risk orderings across admissions even when conditioned on the same discharge-note summary. Thus, representational substitution changed not only how well the model discriminated outcomes overall, but also how admissions were prioritized by predicted risk. The magnitude of these ranking differences is also notable. Kendall’s *τ*-based distances were consistently smaller for comparisons among models trained under the same input configuration with different random seeds than for comparisons across configurations that differed by modality. This pattern suggests that the observed ranking differences are not readily explained by ordinary finetuning variability alone. In the radiology-only setting (RR vs. CXR), *τ*-based distances between report and CXR models were substantially larger than the intra-configuration baselines, indicating pronounced disagreement associated with representational substitution. When discharge-note context was shared (DN+RR vs. DN+CXR), the *τ*-based distances decreased overall, consistent with the shared note constraining model behavior to some extent, but they remained clearly above the intra-configuration baselines. Even under this controlled shared-context setting, the choice of augmentation modality therefore induced meaningful changes in admission-level prioritization.

A plausible explanation is that radiology reports, although clinically useful summaries, do not necessarily preserve all image-visible details that contribute to downstream risk stratification. Our visualization analysis offers suggestive, though limited, evidence in this regard: both activation-based and occlusion-based maps tended to highlight peripheral soft tissues and skeletal structures rather than the lung fields that constitute the primary focus of clinical interpretation. This pattern raises the possibility that prognostically relevant information may reside in features not typically emphasized in radiology reports, including degenerative skeletal changes, body habitus indicators, and the presence of medical devices, rather than in the cardiopulmonary findings that reports are primarily designed to capture. Indeed, the radiologist’s review identified several features that were visible on the images but not explicitly documented in the paired reports, consistent with routine reporting practice in which stable, expected, or non-actionable findings may be abbreviated, deprioritized, or left implicit. These findings may help contextualize why expert-written reports can function as clinically effective summaries without necessarily serving as lossless prognostic surrogates for raw images.

From a clinical AI perspective, these findings suggest that representational modality choice can have behaviorally meaningful consequences. If models are used to identify a limited subset of patients for additional monitoring or intervention, disagreement in risk ranking may affect which patients are prioritized. Accordingly, when one modality is used as a practical substitute for another, evaluation should extend beyond average discrimination to include admission-level ranking agreement.

Nevertheless, this study has several limitations. **1. Generalizability**: This study was conducted on a single cohort derived from MIMIC-IV and MIMIC-CXR, which may limit generalizability to other institutions, reporting styles, and imaging practices. **2. Model pretraining**: We finetuned a general-purpose vision–language model rather than a model pretrained specifically for clinical imaging and text, and the absolute magnitude of both discrimination and ranking differences may change under domain-specific pretraining. **3. Input data constraints**: The imaging input was restricted to a single selected pre-discharge CXR per admission, and therefore does not capture longitudinal evolution across serial studies or multiple views. Similarly, we relied on discharge-note summaries rather than full notes for tractability; although this preprocessing was applied consistently across configurations, summarization may remove or compress information in ways that interact with modality augmentation. **4. Interpretability**: The visualization methods used here are not designed to provide causal attribution, and the absence of clearly interpretable patterns in the occlusion-based maps may reflect both methodological limitations and properties of the model itself. Also, the radiologist review provides contextual rather than causal evidence, and was performed on a limited sample intended to contextualize disagreement rather than establish a definitive mechanism.

## 7. Conclusion

We systematically compared raw CXRs and expert-written radiology reports for post-discharge mortality prediction under a shared discharge-note context. Across input configurations, CXR augmentation achieved the strongest discrimination, whereas report augmentation provided limited benefit. Beyond discrimination, we quantified modality-associated behavioral differences using Kendall’s *τ*-based distance on patient risk rankings. Inter-modality comparisons consistently exceeded intra-modality baselines, indicating that representational modality choice can meaningfully affect which patients are prioritized as high risk, even under shared discharge-note context. This approach is novel, and could be used as a general-purpose tool to measure the relative information content of different information modalities conditioned on a prediction target. This could be a valuable tool as multimodal prediction becomes more common and investigators look to trade off prediction accuracy with challenges of working with heterogeneous data types. A post hoc radiologist review further suggested that reports may omit image-visible details, providing a plausible clinical context for why text may not fully preserve prognostic cues present in raw imaging. Overall, these findings suggest that radiology reports may be imperfect prognostic proxies for raw CXRs and that representational substitution should be evaluated not only by discrimination but also by agreement in risk prioritization across admissions.

## Data Availability

All data analyzed in this study are publicly available from the following sources:
LCD Benchmark: https://github.com/Machine-Learning-for-Medical-Language/long-clinical-doc
MIMIC-IV: https://physionet.org/content/mimiciv/
MIMIC-CXR: https://physionet.org/content/mimic-cxr/
MIMIC-CXR-JPG: https://physionet.org/content/mimic-cxr-jpg/
All datasets are fully de-identified and available to credentialed researchers who complete the required data use agreement on PhysioNet.
No new patient data were collected for this study.
Data processing code will be open-sourced soon.

https://physionet.org/content/mimiciv/

https://physionet.org/content/mimic-cxr/

https://physionet.org/content/mimic-cxr-jpg/

## CRediT authorship contribution statement

C. Kim, W. Yoon, and T. Miller conceived and designed the study. C. Kim and H. Lee performed the experiments, curated the data, and conducted all preprocessing and analyses. H. Lee prepared the figures, and J. Lee manually reviewed discrepancies between radiology reports and chest radiographs. M. Afshar provided clinical expertise on imaging practices and discharge note content. C. Kim drafted the original manuscript, which was critically reviewed and edited by all other co-authors. All authors read and approved the final manuscript.

## Data and code availability

The data underlying this article are available from the LCD benchmark: https://github.com/Machine-Learning-for-Medical-Language/long-clinical-doc. The MIMIC-CXR and MIMIC-CXR-JPG datasets are available via PhysioNet (https://physionet.org/content/mimic-cxr/2.1.0/; https://physionet.org/content/mimic-cxr-jpg/2.1.0/), subject to the applicable data use agreements and credentialed access requirements. The preprocessing and training code will be made publicly available upon acceptance of the paper.

## Ethical statement

This study utilized data from MIMIC-IV database (Johnson et al., 2023, 2019, 2024), including de-identified discharge note, CXRs, and radiology reports. All data have undergone rigorous de-identification in accordance with the HIPAA Safe Harbor standards, as documented in Johnson et al. (2023). In all documents, protected health information (PHI) was removed through date shifting, name redaction, and free-text masking, so no explicit PHI remains; temporal fields (e.g., admission, discharge, and birth or death dates) were uniformly shifted to future years (2100–2200) while preserving relative temporal intervals. In certain experimental settings, we incorporated patient age and race information by referencing the MIMIC-IV mapping tables, which were also used to link CXRs with their corresponding radiology reports and discharge note. However, all data processing and analysis procedures were conducted strictly for research purposes, in full compliance with the MIMIC data use agreement and applicable ethical standards, with no attempt made to re-identify any individual patient.

## Acknowledgements

This work was partially supported by the Korean Institute for Advancement of Technology (KIAT) grant RS2024-00435997. Research reported in this publication was also supported by the National Library of Medicine and National Institute of General Medical Sciences of the National Institutes of Health under Award Numbers R01LM012973 and R01GM114355. The content is solely the responsibility of the authors and does not necessarily represent the official views of theNational Institutes of Health. Additional support was provided by the Ministry of Science and ICT (NRF-2023R1A2C3004176), the Ministry of Health and Welfare of South Korea (HR20C002103), and the Seoul National University Hospital with support from the Ministry of Science and ICT (RS-2023-00262002).

## A. Baseline model selection

We evaluated three candidate vision–language models to identify a base model suitable for capturing clinical signals from discharge notes and radiographic inputs. We used the Huggingface library (Wolf, Debut, Sanh, Chaumond, Delangue, Moi, Cistac, Rault, Louf, Funtowicz, Davison, Shleifer, von Platen, Ma, Jernite, Plu, Xu, Le Scao, Gugger, Drame, Lhoest and Rush, 2020) to load the pretrained weights and conduct the evaluations. Among these candidates, Llama-3.2-11B-Vision-Instruct achieved the highest zero-shot F1 score (Table A.1) and was therefore selected as the baseline VLM for subsequent experiments. For consistency across candidate VLMs, the zero-shot comparison was performed without an additional classification head, requiring each model to directly generate a binary prediction (0: alive, 1: dead). Because calibrated probability estimates were not available in this setting, we used the F1 score as the comparison metric and fixed the temperature at 0 to ensure deterministic outputs. Notably, zero-shot performance decreased when CXRs were added without task-specific fine-tuning, suggesting these models may not reliably extract prognostic information from radiographic inputs.

**Table A.1.** score of baseline candidate models. DN: discharge note summary; CXR: chest radiograph.

| Model | DN | DN+CXR |
| --- | --- | --- |
| Llama-3.2-11B-Vision-Instruct | **0.3502** | **0.3389** |
| Phi-3.5-vision-instruct | 0.1809 | 0.1479 |
| Pixtral-12B | 0.1005 | 0.0989 |

## B. Log-odds Shifts Across Summary Variants and Modalities

Each point represents one admission, and colors indicate the ground-truth label (red: deceased, blue: survived). Axes show the predicted log-odds for the positive class (death) from two paired model configurations. The dashed diagonal denotes equal predicted log-odds between configurations; points above the diagonal indicate higher predicted mortality risk under the configuration on the *y*-axis. Figure B.1 compares admission-level mortality predictions across input configurations on the log-odds scale, where points above the diagonal indicate higher predicted mortality risk under the configuration on the *y*-axis than under the configuration on the *x*-axis. Relative to the text-only settings (DN and DN+RR), incorporating CXRs (DN+CXR) shifts predictions upward for many admissions with death outcomes and downward for many admissions with survival outcomes. By contrast, adding radiology reports (DN+RR) relative to DN tends to increase predicted log-odds more broadly, with many points lying above the diagonal. Overall, these patterns suggest that adding CXRs changes admission-level risk predictions in a more outcome-aligned manner than adding radiology reports.

**Figure B.1.**
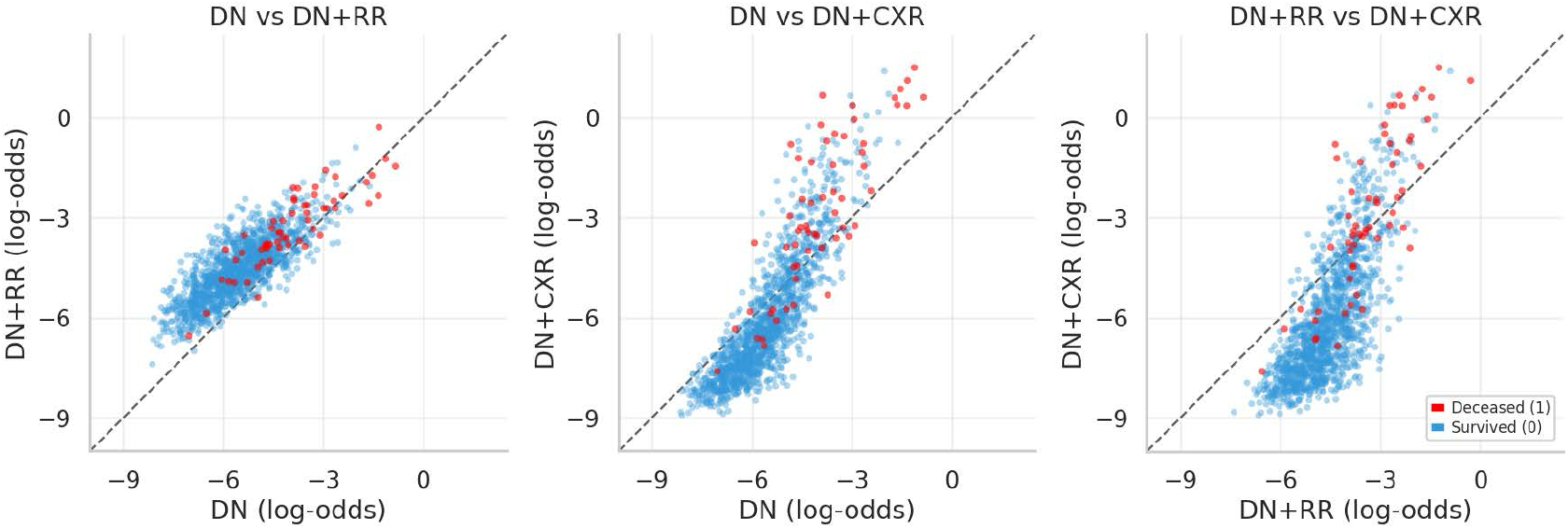
Log-odds scatter plots comparing admission-level mortality predictions across input configurations.

## C. Prompts

### C.1. Summarization Prompt

~~~
System Prompt:
You are a highly trained medical assistant specializing in clinical documentation and summarization. Your task is to produce concise, accurate
  summaries of discharge notes, relying only on the information explicitly provided in the original document. Omit any personally
  identifiable details (e.g., names, ages, races, or ID numbers), and focus strictly on clinically relevant information.
User Prompt:
Here is the clinical discharge document: {DISCHARGE NOTE} Please generate a summary of overall ICU discharge note that includes important
 clinical information. Write the summary as concise, clear, and well-organized bullet points.
~~~

### C.2. Inference Prompt

~~~
Discharge note only
System Prompt: Below is a clinical discharge document. Based on the given clinical context, assess how likely the patient’s out-of-hospital
  mortality is within 30 days.
User Prompt: Here is the clinical discharge document: {DISCHARGE NOTE} Based on the clinical information provided, how likely is the patient’s
  out-of-hospital mortality within 30 days? Please do not explain the reason and respond with one word only: 0:alive, 1:death. Assistant:
CXR only
System Prompt: A single, most recent chest radiograph (CXR) from the patient is provided. Based on the provided CXR, assess how likely the
  patient’s out-of-hospital mortality is within 30 days.
User Prompt: {CXR} Based on the provided single CXR, how likely is the patient’s out-of-hospital mortality within 30 days? Please do not explain
  the reason and respond with one word only: 0:alive, 1:death. Assistant :
Radiology report only
System Prompt: A most recent radiology report from the patient is provided. Based on the provided radiology report, assess how likely the
patient’s out-of-hospital mortality is within 30 days.
User Prompt: Here is the radiology report: {RADIOLOGY REPORT} Based on the radiology report provided, how likely is the patient’s
out-of-hospital mortality within 30 days? Please do not explain the reason and respond with one word only: 0:alive, 1:death. Assistant:
Discharge note + Radiology report
System Prompt: A clinical document and a most recent radiology report are provided. Based on the clinical context and the radiology report,
assess how likely the patient’s out-of-hospital mortality is within 30 days.
User Prompt: Here is the clinical discharge document: {DISCHARGE NOTE} Here is the radiology report: {RADIOLOGY REPORT} Based on the provided
clinical information and radiology report, how likely is the patient’s out-of-hospital mortality within 30 days? Please do not explain the
reason and respond with one word only: 0:alive, 1:death. Assistant:
Discharge note + CXR
System Prompt: A clinical document and a single, most recent chest radiograph (CXR) from the patient are provided. Based on the clinical context
  and the provided CXR, assess how likely the patient’s out-of-hospital mortality is within 30 days.
User Prompt: {CXR} Here is the clinical discharge document: {DISCHARGE NOTE} Based on the provided clinical information and single CXR, how
  likely is the patient’s out-of-hospital mortality within 30 days? Please do not explain the reason and respond with one word only:
  0:alive, 1:death. Assistant:
~~~

## D. Dataset statistics

**Figure D.1.**
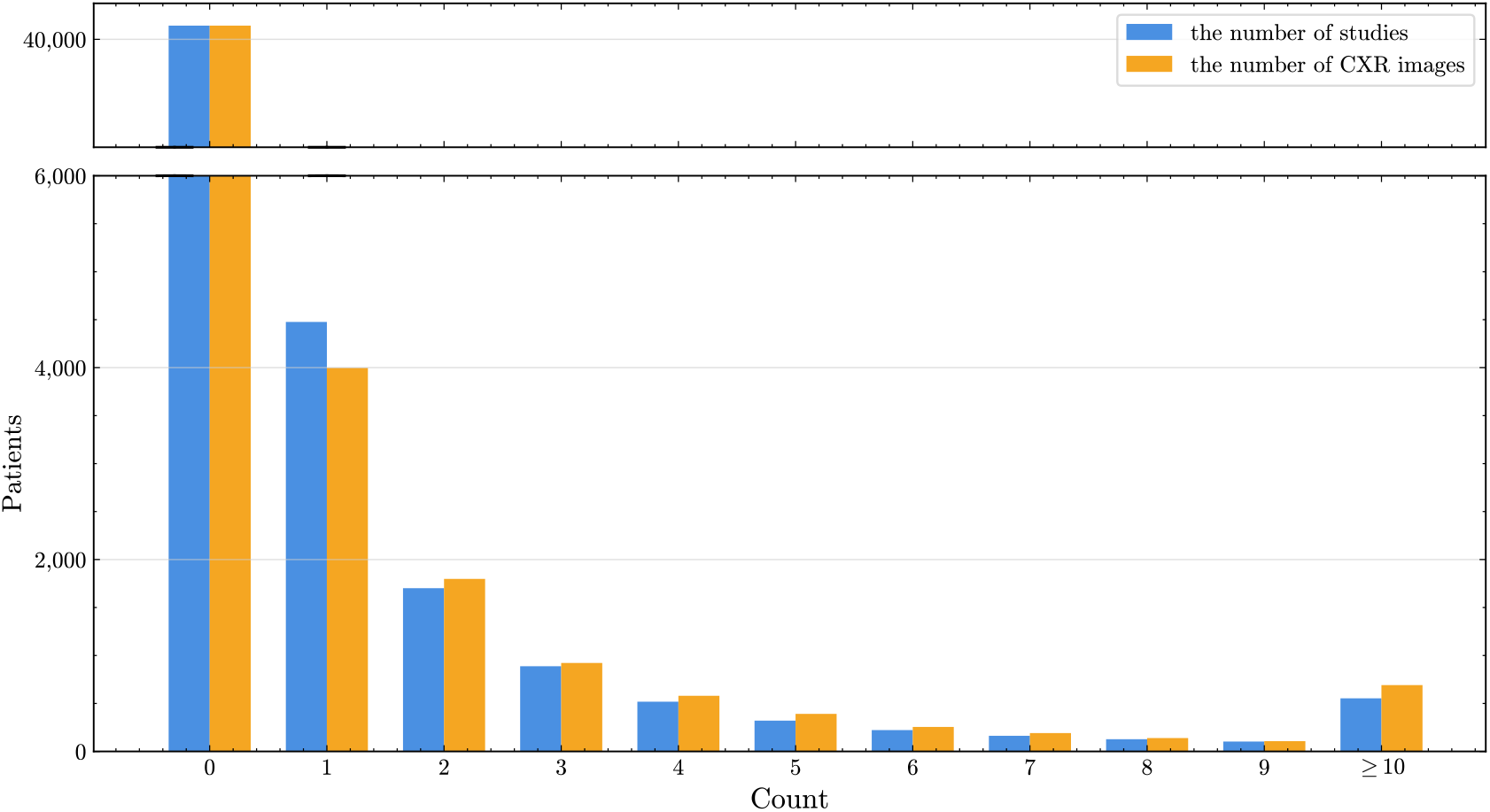
Distribution of admissions by the number of radiographic studies and the number of chest radiograph (CXR) images acquired during the ICU stay. Because a single study may include multiple images, we report these two counts separately for each patient. The figure presents aggregated counts across the Train, Dev, and Test splits, which showed similar overall distributions.

## E. Token lengths

**Figure E.1.**
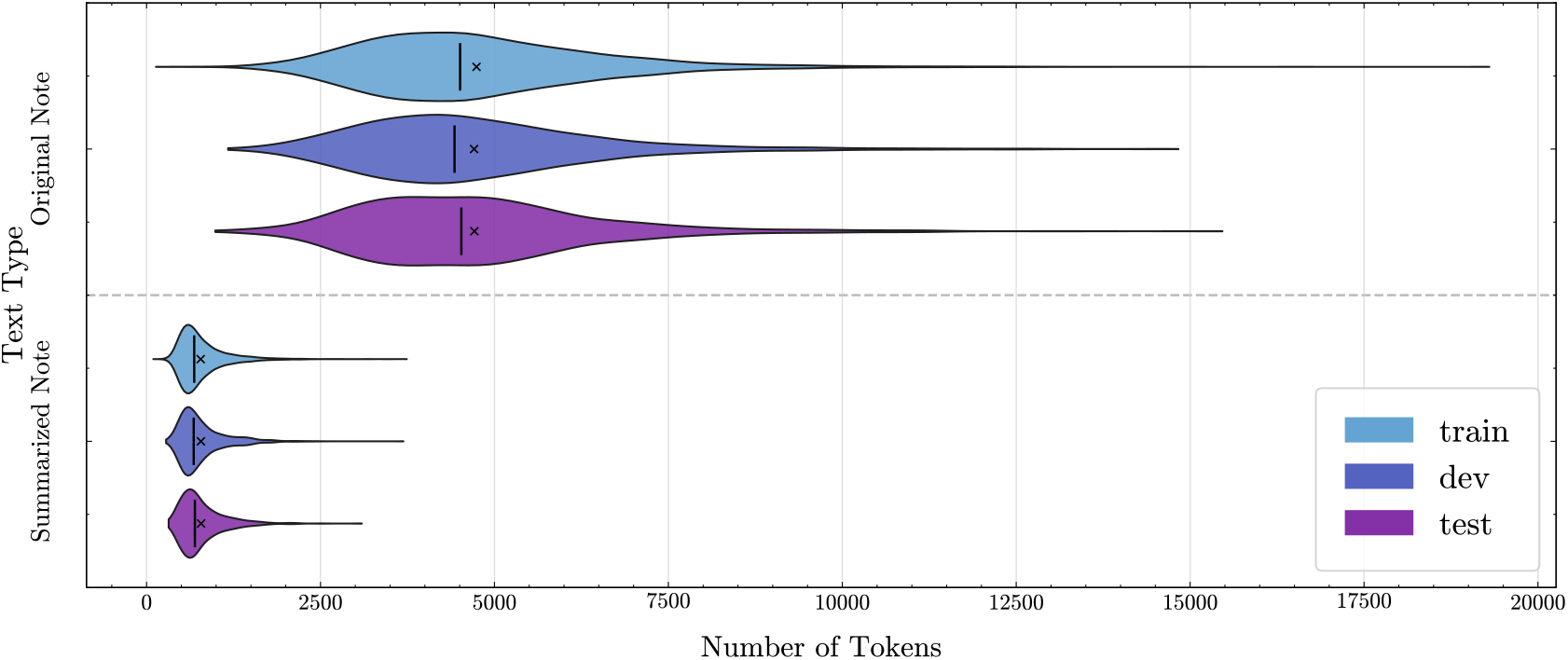
Distribution of the number of total tokens for the original discharge notes and their summaries.

## F. Training hyperparameters

**Table F.1.** Hyperparameter search space and selected values for each model and input configuration. DN = discharge note, RR = radiology report, CXRs = chest radiographs.

| Hyperparameter | Search space | DN | DN+RR | DN+CXR |
| --- | --- | --- | --- | --- |
| **SVM** |  |  |  |  |
| Kernel | [ linear, rbf ] | linear | linear | – |
| C | [ 0.1, 1, 10 ] | 1 | 1 | – |
| Feature dimension | [5000] | 5000 | 5000 | – |
| **ModernBERT** |  |  |  |  |
| Learning rate | [ 2e-5, 1e-5, 5e-6 ] | 1e-5 | 1e-5 | – |
| Batch size | [ 8, 16 ] | 8 | 8 | – |
| Epochs | [ 10, 20, 30 ] | 20 | 20 | – |
| **LLaMA 3.2-11B-vision-instruct** |  |  |  |  |
| Learning rate | [ 2e-5, 1e-5, 5e-6, 2e-6, 1e-6 ] | 1e-6 | 2e-6 | 2e-6 |
| Batch size | [ 8, 16 ] | 16 | 16 | 16 |
| Epochs | [ 10, 20, 30 ] | 30 | 30 | 30 |

## G. CXR Selection Decision Tree

**Figure G.1.**
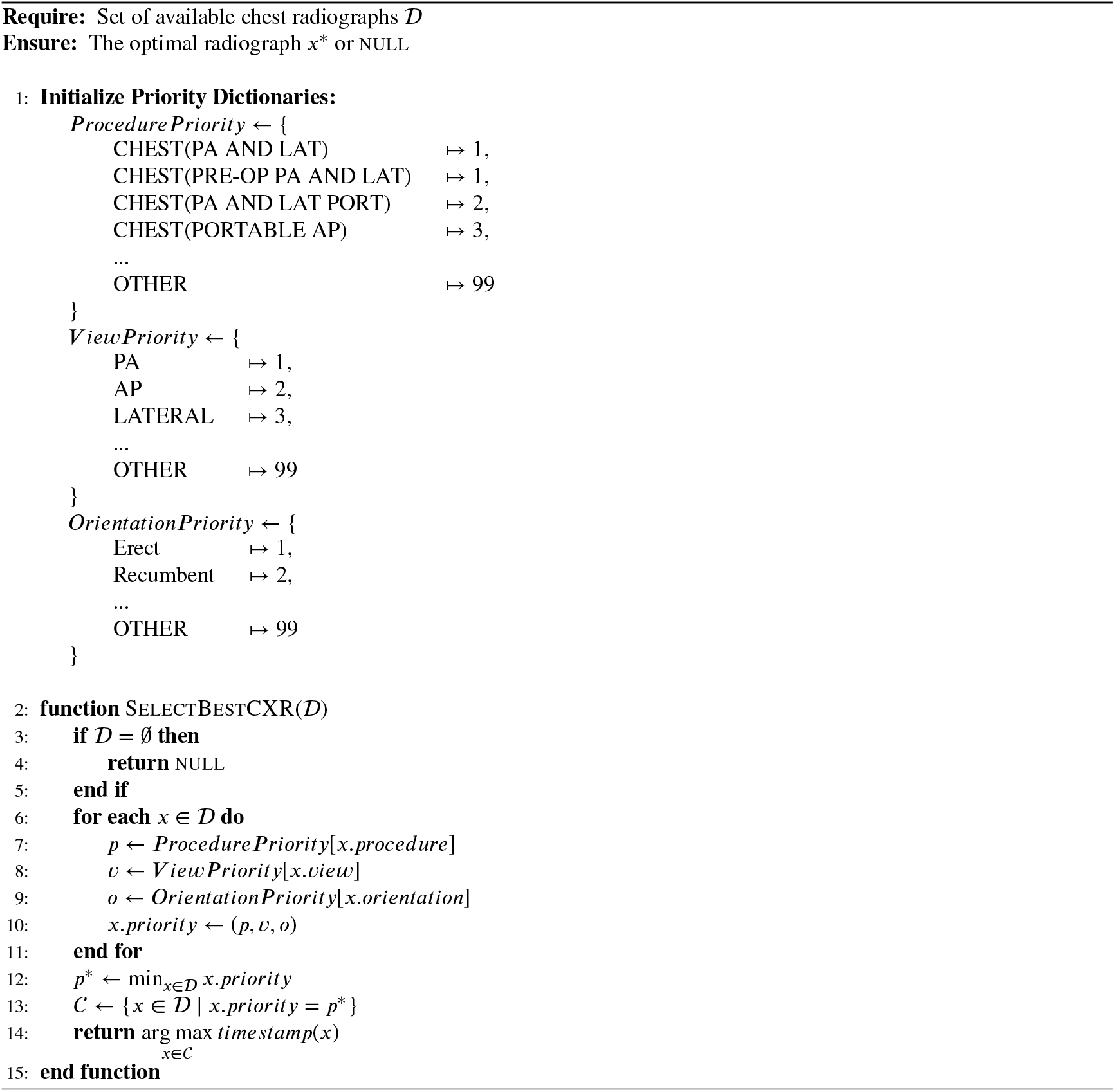
Selection algorithm for optimal chest radiographs.

